# Can eraDOCator-60 Decrease MDRs and HAIs? An Evaluation of the Efficacy of eraDOCator-60 in a Randomized Clinical Trial in a Community Hospital

**DOI:** 10.1101/2023.05.26.23290573

**Authors:** Martin Johns, Matthew Machata, Stephanie Liseno, Joseph del Castillo

## Abstract

**Background:** There is a significant transmission of contaminants in the healthcare setting. Daily disinfection utilizing ammonium and chlorine-based products can lead to adverse health effects such as asthma, cancer, and other serious health issues.

**Methods:** This study evaluated the effectiveness of eraDOCator-60 in a health care facility. This randomized trial took place at Copley Hospital in Morristown, Vermont. Separate areas of the hospital were cleaned and disinfected in one step with eraDOCator-60. A Charm analyzer was utilized to evaluate the efficacy of disinfection before and after 1 minute application of eraDOCator-60. The Charm analyzer detects Adenosine Triphosphate (ATP) presence measured in Relative Light Units (RLUs).

**Results:** The median number of RLUs decreased from 52,874 s to 0 RLUs after one-minute eraDOCator-60 dwell time in the emergency room; 18.611 RLUs to 0 RLUs in the medical-surgical unit, 41,507 RLUs to 0 RLUs in the cafeteria; 24,932 RLUs to 0 RLUs in the birthing center.

**Conclusions:** EraDOCator-60 reduced contamination levels on all surfaces in the acute care setting down to a value of zero following a 1-minute dwell time in less than 5% soil load.

## INTRODUCTION

The U.S. Centers for Disease Control and Prevention (CDC) estimates that 3–4 percent of all hospital admissions result in a healthcare-associated infection (HAIs) [1], culminating in approximately 687,200 infections and 72,000 deaths each year as well as $28–45 billion in excess costs. [1–4] It is not uncommon to find acute care centers using multiple products to clean and disinfect surfaces. These products, typically quaternary ammonium-based, vary in their dwell time to kill bacteria, and have potential carcinogenic and harmful properties (**Table 1**). [5–9] Quaternary ammonium and Chloride based chemicals, for example, also leave sublethal residues that lead to multi drug resistant (MDRs) bacteria such as MRSA and VRE, which also significantly increases the risk of acquiring an HAI [5]. These products have been found to disable equipment due to corrosion, strip and erode floors, harm the environment, and harm people who come into contact with these chemicals on a regular basis. [6,7,10].

**Table 1.** Hazardous properties of common disinfectants

|  | **Inorganic alcohols**<br>**(e.g. phenols or**<br>**thymols)** | **Chlorine**<br>**products (e.g.**<br>**bleach)** | **Ammonium**<br>**products** | **eraDOCator 60**<br>**and 120** |
| --- | --- | --- | --- | --- |
| Has inorganic active components | ✓ | ✓ | ✓ |  |
| Leaves toxic byproducts | ✓ | ✓ | ✓ |  |
| Produces toxic gas | ✓ | ✓ | ✓ |  |
| Skin irritant | ✓ | ✓ | ✓ |  |
| Skin pigmentation changes | ✓ | ✓ |  |  |
| Increases cancer risk | ✓ | ✓ | ✓ |  |
| Can create water toxicity | ✓ | ✓ | ✓ |  |
| Contributes to ozone depletion |  | ✓ |  |  |
| Affects aquatic life/coral reefs |  | ✓ | ✓ |  |
| COPD, bronchitis, asthma and rhinitis | ✓ | ✓ | ✓ |  |
| Chronic chemical pneumonitis |  | ✓ | ✓ |  |
| Fetal damage or reproductive issues | ✓ |  | ✓ |  |
| Dental cavities/dental loss |  | ✓ |  |  |

EraDOCator-60, a peracetic acid (PAA) based broad-spectrum disinfectant, was designed to clean, disinfect, and sanitize hard non-porous surfaces in one minute. Due to its ability to kill bacteria, viruses, and fungi in a food safe manner, it is intended to act as a multi-purpose disinfectant, simplifying protocols for staff and creating safe environments in the acute care setting. It works as a potent microbe oxidizer and is classified as a broad-spectrum sterilizer. The PAA diluted product is the safest broad spectrum, hospital-grade cleaner and disinfectant available for both humans and the environment. Following disinfection, the product evaporates into carbon dioxide and water, leaving no residue on any surface which reduces the instances of both MDRs and HAIs.

The purpose of this study was to quantify the multi-surface disinfecting properties of eraDOCator-60 in a hospital setting.

## METHODS

The primary objective was to analyze the presence and reduction of ATP contained on hospital surfaces that were cleaned and sanitized with eraDOCator-60. The secondary objective was to show during the cleaning process that the product is safe and disinfects surfaces faster than quaternary ammonium cleaning products, which require an average dwell time of three to ten minutes.

### Study Design and Setting

The testing of eraDOCator-60 took place in Copley Hospital in Morristown, Vermont. Copley Hospital is a community hospital/critical access hospital that incorporates and specializes in Emergency Medicine, Orthopedics, General Medicine, and Obstetrics. Test locations were identified based on the most commonly utilized areas for patients and staff that were also in need of a fast paced turnover clean in the day-to-day flow of the hospital. Multiple areas of the hospital, including the birthing center and the cafeteria, were chosen for the study.

The level of surface contamination was tested with a Charm Analyzer that detects levels of ATP in RLUs. “ATP is a high-energy molecule that is used by living cells as their primary source of energy. Animal, plant, bacteria, yeast, and mold break down ATP to drive several biological processes. ATP may be cellular (e.g., within viable or dead cells) and extra-cellular (free ATP).[13]” When measuring ATP levels in RLU’s there is no way to differentiate between bacterial cells and non-bacterial cells because ATP is present in all biological material. However, the increased incidence of ATP on a surface indicates that there is a higher likelihood of bacterial presence or there is potential for rapid bacterial growth. Essentially, higher RLU numbers indicate high amounts of ATP and a more contaminated surface. For a surface ATP test, a passing RLU is 0–50, and a failure is anything greater than 50. This was correlated with Agar plate testing (< 50 RLUs showed nearly no growth on Agar plates) see figure 2.

**Figure 1.**
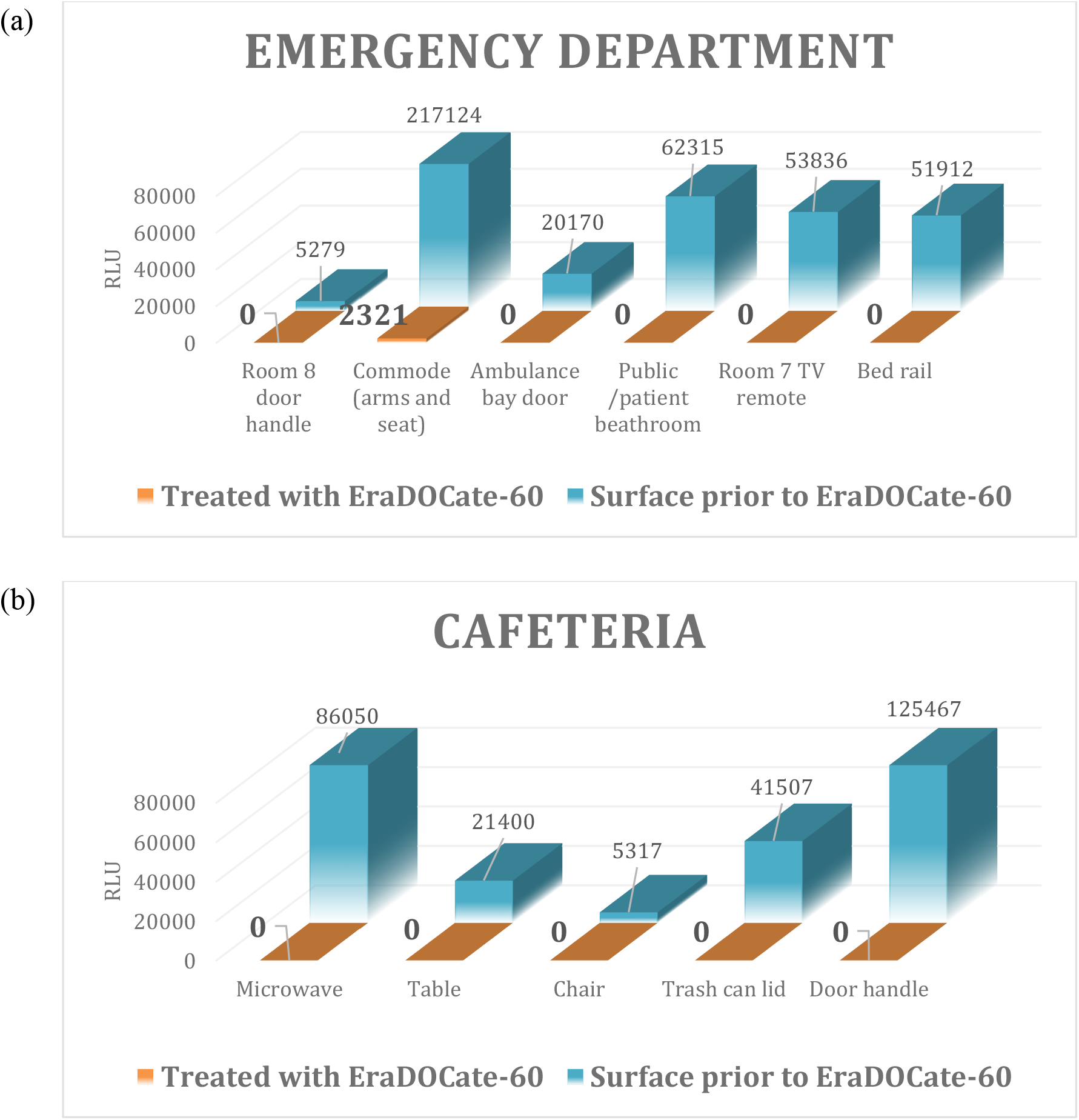
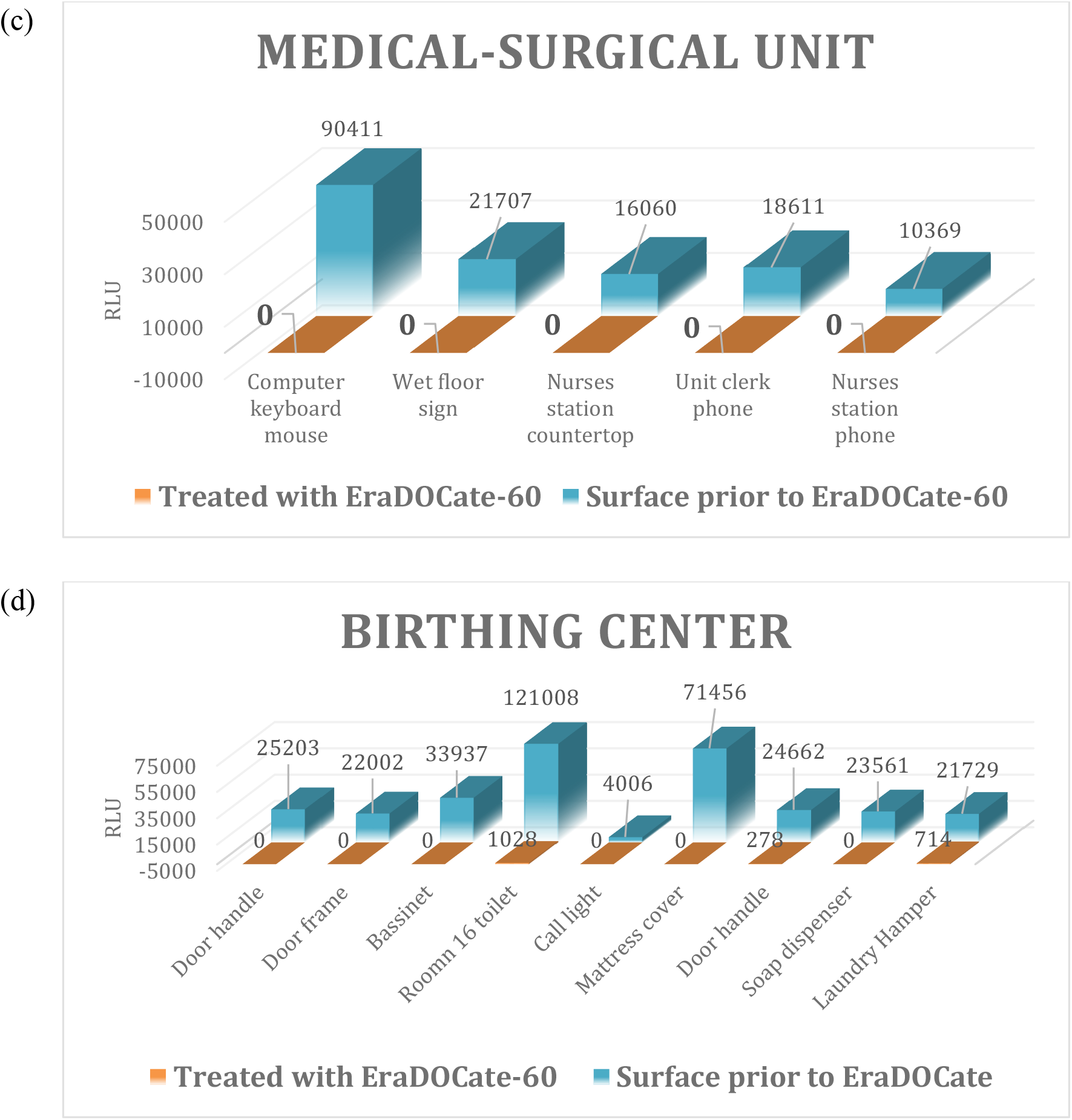
RLU levels on Emergency room surfaces (a); Cafeteria surfaces (b); Medical-surgical unit surfaces (c); Birthing center surfaces (d) before and after 1-minute eraDOCator-60 disinfection.

**Figure 2.**
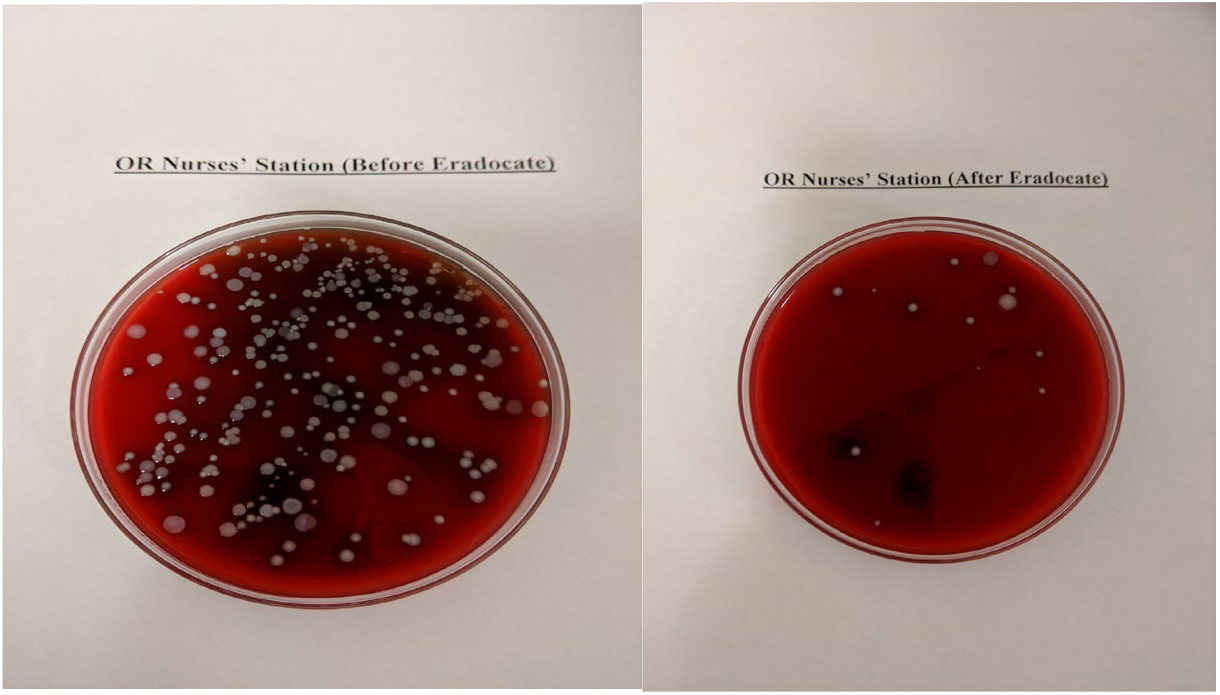
Nurses station contamination quantities before (top) and after (below) eraDOCator-60 disinfection.

### Study Protocol

EraDOCator-60 was applied to the surfaces in pre-specified hospital areas. Surfaces in the identified areas below were swabbed prior to and after a one-minute application of the product by a wet towel. Areas with higher than a 5% soil load required organic removal prior to disinfection. The cleaning of the surfaces was followed per the eraDOCator-60 instructions.

## RESULTS

Contaminants were found on the surface of each area of the hospital that were tested prior to the eraDOCator-60 disinfection. The areas occupied by both patients and staff are outlined below.

### Emergency Room

The Emergency room consisted of swabs on a commode arm/seat, ambulance bay door, public/patient bathroom, TV remote, and bed rail. The median number of RLUs for the Emergency department was 52,874 RLUs prior to the eraDOCator-60 disinfection. There was contaminate found on every surface. Swabs came from the door handle (5,279 RLUs), the commode (217,124 RLUs), the ambulance bay door (20,170 RLUs), the public/patient bathroom (62,315 RLUs), the TV remote (53,836 RLUs), and the bed rail (51,912 RLUs).

Following the eraDOCator-60 disinfection clean after one minute, only the commode had a >0 RLU reading (2321 RLU). Following a second wipe (two-minute dwell time) all results equaled 0 RLUs (**Figure 1A**). [The commode had a visible > 5% feces soil burden]

### Cafeteria

The total median number of RLUs in the cafeteria equaled 41,507 RLUs. Swabs came from a microwave (86,050 RLUs,) a table (21,400 RLUs), a chair (5,317 RLUs), a trash can lid (41,507 RLUs), and the door handle (125,467 RLUs). Following eraDOCator-60 disinfection with a one-minute dwell time, each follow up swab was 0 RLUs (**Figure 1B**).

### Medical-Surgical Unit

The median number of RLUs measured in the Medical Surgical Unit equaled 18,611 RLUs

. Swabs came from a computer keyboard and mouse (90,411 RLUs), a wet floor sign (21,707 RLUs), the nurses station countertop (16,060 RLUs), the unit clerk phone (18,611 RLUs), and the nurses station phone (10,369 RLUs). Following eraDOCator-60 disinfection with a dwell time of one minute, all swabs were 0 RLUs (**Figure 1C and 2**).

### Birthing Center

The median number of RLUs in the Birthing Center prior to eraDOCator-60 disinfection was 24,932 RLUs. Swabs came from the bassinet (205,004 RLUs), the door handle (25,203 RLUs), the door frame (22,002 RLUs), a second bassinet (36,937 RLUs), a patient toilet (121,008 RLUs), a call light (4,006 RLUs), a mattress cover (71,456 RLUs) a door handle (24,662 RLUs) a soap dispenser (23,561 RLUs), and the laundry hamper (21,729 RLUs).

Following a one-minute dwell time of eraDOCator-60, the median number of RLUs totaled 714 RLUs, while following a second minute dwell time, each test was in 0 RLUs (**Figure 1D**). [The Birthing Center had a visible coating of lochia and blood greater than 5%. which required initial eraDOCator-60 soil removal (clean) prior to eraDOCator-60 disinfecting]

### Operating Room

The cabinet of the Operating Room measured 75 RLUs prior to eraDOCator-60 disinfection and measured 4 RLUs following the disinfection using eraDOCator-60.

### Questionnaire Results

In addition to eraDOCator-60 surface disinfection results, we surveyed the Environmental Staff on their first-hand experience cleaning with the product. We received 17 responses from the staff concerning the safety and efficacy of eraDOCator-60. When asked if eraDOCator-60 meets the needs of the Environmental Service Workers, all 17 (100%) respondents answered yes. When asked if the Environmental Service workers were able to implement eraDOCator-60 in the facility in a timely manner, 16 (94%) respondents answered yes, and 1 (6%) respondent skipped the question. The workers were asked if eraDOCator-60 was “Easy to use.” All 17 (100%) respondents of the survey answered yes.

## DISCUSSION

The rise in HAIs and MDRs has led to an increasing focus on hospital disinfection procedures. However, concerns over exposure to irritant and toxic chemicals found in common cleaning products has led to a need for non-toxic alternatives to avoid respiratory illnesses, headaches, and eventual cancers and long term disability [5-10]. The increasing complexity of hospital disinfection procedures, often requiring ≥10-minute dwell times and multiple disinfectants in the same space led to concerns about compliance and cleaning effectiveness. EraDOCator-60, a product derived from vinegar and hydrogen peroxide, was designed as a one-step cleaner and disinfectant with a dwell time of one minute for use in a hospital setting. The product allows for a rapid turnaround time in disinfection of areas between patients (such as ER and OR).

In this study, eraDOCator-60 was found to disinfect all non-porous hard surfaces in the acute care setting with a one-minute dwell time to a value of zero RLUs. In instances in greater than a 5% soil burden (such as a large amount of blood), a separate cleaning needs to take place, This study proves the efficacy of the product eraDOCator-60 as a potent disinfectant while leaving no residues leading to reduced MDRs and HAIs. For the two years of 100 percent implementation of eraDOCator-60, Copley Hospital has had zero incidence of HAIs, EraDOCator-60 enables environmental service workers to employ the safest available, non-toxic, water and food safe product for one step cleaning and disinfecting. As the study illustrates, eraDOCator-60 brings levels of ATP to zero RLUs, indicating that the ATP present is not sufficient to harbor bacterial growth. This makes the product perfect for all areas of acute care, in places where sanitation is imperative, as is rapid room turnaround time. Copley Hospital has been using eraDOCator-60 throughout the facility for two years, and it has replaced more than 90 percent of the cleaning chemicals in the hospital.

## CONCLUSIONS

Most products used in the acute care setting daily, not only cause health issues, but leave residues on surfaces, degrade medical equipment, and are extremely costly to healthcare facilities. These residues are the birthplace of MDRs and HAIs. Utilizing eraDOCator-60 enables the replacement of over 90 percent of harmful substances in the workplace, reducing exposure of staff and patients to toxic chemicals. With a base of PAA and hydrogen peroxide, the one-step cleaner/disinfectant is food and water safe, and is currently certified by the EPA, FDA, and CDC.

EraDOCate’s closed loop dilution system ensures the safety of workers and ease of use throughout the acute care setting. It decreases waste using a one-product model and is eco-friendly. EraDOCate’s straightforward implementation into the facility, ease of use of eraDOCator-60, and ability to meet the needs of staff using the product each day outlines its commercial effectiveness. This allows for timely turnover between patients, prevention of MDRs and HAIs, and ensures safety of people and the environment. Innovation in efficiency, satisfaction of staff, and having an eco-friendly one product model delivers health, wellness, plus huge cost savings for healthcare systems.

## Data Availability

All data produced in the present study are available upon reasonable request to the authors
All data produced in the present work are contained in the manuscript

## ACKNOWLEDGEMENTS

Medical writing and editorial support was provided by Claire L Jarvis, PhD. Background and site support was provided by Kathryn Bittner, RN, BSN, MPH.

